# Characterization of Microbial Co-infections in the Respiratory Tract of hospitalized COVID-19 patients

**DOI:** 10.1101/2020.07.02.20143032

**Authors:** Huanzi Zhong, Yanqun Wang, Zhun Shi, Lu Zhang, Huahui Ren, Weiqun He, Zhaoyong Zhang, Airu Zhu, Jingxian Zhao, Fei Xiao, Fangming Yang, Tianzhu Liang, Feng Ye, Bei Zhong, Shicong Ruan, Mian Gan, Jiahui Zhu, Fang Li, Fuqiang Li, Daxi Wang, Jiandong Li, Peidi Ren, Shida Zhu, Huanming Yang, Jian Wang, Karsten Kristiansen, Hein Min Tun, Weijun Chen, Nanshan Zhong, Xun Xu, Yi-min Li, Junhua Li, Jincun Zhao

**Affiliations:** BGI-Shenzhen, Shenzhen, 518083, China; State Key Laboratory of Respiratory Disease, National Clinical Research Center for Respiratory Disease, Guangzhou Institute of Respiratory Health, the First Affiliated Hospital of Guangzhou Medical University, Guangzhou, Guangdong, 510120, China; Laboratory of Genomics and Molecular Biomedicine, Department of Biology, University of Copenhagen, 2100 Copenhagen, Denmark; Institute of Infectious disease, Guangzhou Eighth People’s Hospital of Guangzhou Medical University, Guangzhou, Guangdong, 510060, China; Department of Infectious Diseases, Guangdong Provincial Key Laboratory of Biomedical Imaging, Guangdong Provincial Engineering Research Center of Molecular Imaging, The Fifth Affiliated Hospital, Sun Yat-sen University, Zhuhai, Guangdong, 519000, China; School of Future Technology, University of Chinese Academy of Sciences, Beijing 101408, China; Shenzhen Key Laboratory of Unknown Pathogen Identification, BGI-Shenzhen, Shenzhen, 518083, China; The Sixth Affiliated Hospital of Guangzhou Medical University, Qingyuan People’s Hospital, Qingyuan, Guangdong, China; Yangjiang People’s Hospital, Yangjiang, Guangdong, China; State Key Laboratory of Bioelectronics, School of Biological Science and Medical Engineering, Southeast University, Nanjing 210096, China; Guangdong Provincial Key Laboratory of Human Disease Genomics, Shenzhen Key Laboratory of Genomics, BGI-Shenzhen, Shenzhen 518083, China; BGI Education Center, University of Chinese Academy of Sciences, Shenzhen, 518083, China; Shenzhen Engineering Laboratory for Innovative Molecular Diagnostics, BGI-Shenzhen, Shenzhen, 518120, China; James D. Watson Institute of Genome Science, Hangzhou, 310008, China; Guangdong Provincial Academician Workstation of BGI Synthetic Genomics, BGI-Shenzhen, Shenzhen, 518120, China; HKU-Pasteur Research Pole, School of Public Health, Li Ka Shing Faculty of Medicine, The University of Hong Kong, Hong Kong SAR, China; BGI PathoGenesis Pharmaceutical Technology Co., Ltd, BGI-Shenzhen, Shenzhen 518083, China; Guangdong Provincial Key Laboratory of Genome Read and Write, BGI-Shenzhen, Shenzhen, 518120, China; School of Biology and Biological Engineering, South China University of Technology, Guangzhou, China

## Abstract

**Background:** Severe acute respiratory syndrome coronavirus 2 (SARS-CoV-2) has caused a global pandemic of Coronavirus disease 2019 (COVID-19). However, microbial composition of the respiratory tract and other infected tissues, as well as their possible pathogenic contributions to varying degrees of disease severity in COVID-19 patients remain unclear.

**Method:** Between January 27 and February 26, 2020, serial clinical specimens (sputum, nasal and throat swab, anal swab and feces) were collected from a cohort of hospitalized COVID-19 patients, including 8 mildly and 15 severely ill patients (requiring ICU admission and mechanical ventilation), in the Guangdong province, China. Total RNA was extracted and ultra-deep metatranscriptomic sequencing was performed in combination with laboratory diagnostic assays. Co-infection rates, the prevalence and abundance of microbial communities in these COVID-19 patients were determined.

**Findings:** Notably, respiratory microbial co-infections were exclusively found in 84.6% of severely ill patients (11/13), among which viral and bacterial co-infections were detected by sequencing in 30.8% (4/13) and 69.2% (9/13) of the patients, respectively. In addition, for 23.1% (3/13) of the patients, bacterial co-infections with *Burkholderia cepacia* complex (BCC) and *Staphylococcus epidermidis* were also confirmed by bacterial culture. Further, a time-dependent, secondary infection of *B. cenocepacia* with expressions of multiple virulence genes in one severely ill patient was demonstrated, which might be the primary cause of his disease deterioration and death one month after ICU admission.

**Interpretation:** Our findings identified distinct patterns of co-infections with SARS-CoV-2 and various respiratory pathogenic microbes in hospitalized COVID-19 patients in relation to disease severity. Detection and tracking of BCC-associated nosocomial infections are recommended to improve the pre-emptive treatment regimen and reduce fatal outcomes of hospitalized patients infected with SARS-CoV-2.

**Funding:** National Science and Technology Major Project of China, National Major Project for Control and Prevention of Infectious Disease in China, the emergency grants for prevention and control of SARS-CoV-2 of Ministry of Science and Technology and Guangdong province, Guangdong Provincial Key Laboratory of Genome Read and Write, Guangdong Provincial Academician Workstation of BGI Synthetic Genomics, and Shenzhen Engineering Laboratory for Innovative Molecular Diagnostics.

## Introduction

As of July 1^st^, 2020, severe acute respiratory syndrome coronavirus 2 (SARS-CoV-2) has infected over 10 million and resulted in more than 500 thousand deaths worldwide^1^. The pandemic poses a significant threat to public health and global economy.

Respiratory viruses, such as coronaviruses and influenza, can lead to acute damage of the epithelial barrier and facilitate invasions of other pathogens^2,3^. For instance, secondary infections by *Stenotrophomonas maltophilia, Klebsiella pneumoniae* or *Escherichia coli* were reported to cause serious complications in patients with SARS, such as bacteremia, sepsis and nosocomial pneumonia (NP)^4^. In addition, *Streptococcus pneumoniae, Haemophilus influenzae* and *Staphylococcus aureus* were frequently associated with NP and mortality in influenza pandemics^5^. It was estimated that approximately 29%-55% of the total 300,000 deaths in the 2009 H1N1 pandemic were caused by secondary bacterial NP^6–8^.

Concerns about co-infections of SARS-CoV-2 with known viruses, bacteria and fungi have also been raised. In severely ill patients, acute respiratory distress syndrome (ARDS) tends to rapidly deteriorate the conditions of the patient, and mechanical ventilation is generally required^9,10^. Such invasive procedures can further increase the risks of ventilator-associated pneumonia (VAP) in these patients^11^. In 99 confirmed Wuhan patients enrolled in January 2020, one (1%) had positive cultures of *Acinetobacter baumannii, K. pneumoniae* and *Aspergillus flavus*, and four (4%) were diagnosed with infection by *Candida*, but no influenza viruses were detected^9^. A later retrospective study in Wuhan patients further demonstrated that half of the deceased patients (27 out of 54) had experienced secondary infections^12^. By using real-time reverse transcriptase-polymerase chain reaction tests, Kim et al. recently reported a 20.7% (24 out of 116 specimens) co-infection rate with SARS-CoV-2 and other respiratory viruses in Northern California, including rhinovirus (6.9%) and orthopneumovirus (5.2%)^13^. However, microbial co-infections as well as their possible effects on clinical outcomes of SARS-CoV-2 infected patients remain largely unknown.

Here, by simultaneously applied diagnostic technologies (cultures and colorimetric assays) and metatranscriptomic sequencing, microbial co-infections in a Guangdong cohort of 23 patients hospitalized with SARS-CoV-2 infection were comprehensively evaluated. High co-infection rate (84.6%) of at least one type of respiratory pathogens among severely ill patients was documented, whereas none of the patients with mild symptoms presented evidence of co-infection. Our results further demonstrated that BCC bacteria, *Staphylococcus epidermidis* and *Mycoplasma spp*. were the most prevalent pathogenic bacteria associated with culture-confirmed and/or metatranscriptomic-detected co-infections, which might increase risk for prolonged ICU stay or even mortality. Our findings demonstrate the value of metatranscriptomics for unbiased co-detection of respiratory pathogens together with SARS-CoV-2 and provide in-time information and useful practical suggestions regarding the adequate monitoring and management of bacterial co-infections in severely ill COVID-19 patients.

## Methods

### Enrollment of hospitalized patients with SARS-CoV-2 infection, collection of clinical specimens

Twenty-three patients admitted to hospital in the period January 10-March 31, 2020 with confirmed SARS-CoV-2 infection based on a positive SARS-CoV-2 test were included. A total of 67 clinical specimens from the respiratory and gastrointestinal tract (including throat swab, nasal swab, sputum, anal swab and feces) were collected from the above patients at the First Affiliated Hospital of Guangzhou Medical University (thirteen patients), the Fifth Affiliated Hospital of Sun Yat-sen University (two patients), Yangjiang People’s Hospital (five patients) and Qingyuan People’s Hospital (three patients) during the period (between January 27 and February 26, 2020). Patients were classified into mild (n=8, without intensive care unit admission) and severe (n=15, with ICU admission) cases based on their severity of SARS-CoV-2 symptoms. Detailed de-identified information for patients and clinical specimens are presented in **appendix 2 p 2: TableS1** and **appendix 2 p 3: TableS2**, respectively.

### Ethics statement

The study was reviewed and approved by the ethics committees of all the four hospitals and the institutional review board of BGI-Shenzhen.

### Laboratory diagnosis of nosocomial bacterial and fungal infections

All severely ill COVID-19 patients admitted to the ICU for more than 48 hours were monitored for nosocomial infections, which were defined according to the definitions of the US Centers for Disease Control and Prevention (CDC) ^14^. Culturing of sputum and nasal secretions was conducted according to standard protocols for the diagnosis of nosocomial microbial infections (hospital-acquired and ventilator-associated) in all severely ill COVID-19 patients with ICU admission^15^. Blood samples of patients (P01 and P05) hospitalized in the Fifth Affiliated Hospital of Sun Yat-sen University were collected for colorimetric assay-based (1-3)-β-D-glucan test to diagnose fungal infection (Dynamiker Biotechnology, Tianjin, China, Catalogue number: DNK-1401-1).

### RNA extraction, metatranscriptomic library preparation and sequencing

For each sample, total RNA was extracted (QiAamp RNeasy Mini Kit, Qiagen, Germany) and the concentration was quantified (Qubit RNA HS Assay Kit, Thermo Fisher Scientific, Waltham, MA, USA). Purified RNA samples were then subjected to pre-processing, DNA nanoball-based libraries construction and high-throughput metatranscriptomic sequencing on the DNBSEQ-T7 platform (100nt paired-end reads, MGI, Shenzhen, China) as described previously ^16^.

### Identification and removal of human RNA reads from metatranscriptomic data

For each sample, the raw metatranscriptomic reads were processed using Fastp (v0.19.5, default settings) ^17^ to filter low-quality data and adapter contaminations and generate the clean reads for further analyses. Human-derived reads were identified with the following steps: 1) identification of human ribosomal RNA (rRNA) by aligning clean reads to human rRNA sequences (28S, 18S, 5.8S, 45S, 5S, U5 small nuclear RNA, as well as mitochondrial mt12S) using BWA-MEM 0.7.17-r1188 ^18^; 2) identification of human transcripts by mapping reads to the hg19 reference genome using the RNA-seq aligner HISAT2 (vertion 2.1.0, default settings) ^19^; and 3) a second-round identification of human reads by aligning remaining reads to hg 38 using Kraken2 (version 2.0.8-beta, default settings) ^20^. All human RNA reads were then removed to generate qualified non-human RNA-seq data. The number of human RNA-seq reads identified at each step is presented in **appendix 2 p 4: TableS3**.

### Characterization of viral communities in hospitalized patients with SARS-CoV-2 infection

Before identification of virome and microbiota, SortMeRNA version 4.2.0^21^ (default settings) was applied to filter microbial rRNA from non-human metatranscriptomic data. The remaining non-human non-rRNA reads were processed by Kraken2X v2.08 beta (default parameters)^20^ with a self-built viral protein database by extracting protein sequences from all complete viral genomes deposited in the NCBI RefSeq database (8,872 genomes downloaded on March 1st, 2020 including the SARS-CoV-2 complete genome reference sequence, GCF_009858895.2). The number of reads annotated to each viral family was summarized based on the read alignment results of Kraken 2X, and all RNA reads annotated to family *Coronaviridae* were considered as SARS-CoV-2-like reads. For each sample, the ratio of SARS-CoV-2-like reads to total clean reads (referred to the SARS-CoV-2 viral load), and the ratio of SARS-CoV-2-like reads to total viral reads were calculated accordingly (**appendix 2 p 5: TableS4**). After ranking the aligned reads of all detected viral species in each sample, non-*Coronaviridae* viral species with more than 10,000 aligned RNA reads were presented (**appendix 2 p 6: TableS5**) and selected for co-detection with known respiratory viruses in hospitalized patients with SARS-CoV-2 infection. Three human respiratory viral species (including human alphaherpesvirus 1, human orthopneumovirus and rhinovirus B) detected in respiratory samples from 4 severe cases (P01, P05, P09 and P13) met the above criterion. Representative genomes of each species including human herpesvirus 1 strain 17 (NC_001806.2), human orthopneumovirus subgroup A (NC_038235.1) and rhinovirus B isolate 3039 (KF958308.1) were downloaded from NCBI. For each patient, one representative sample (P01N201, P05S207, P09N205 and P12T211) with the highest number of reads assigned to the targeted species was used for coverage analysis. Reads assigned to a given species were aligned against the corresponding reference genome by bowtie2 v2.3.0 (the ‘-sensitive’ mode)^22^. Sequencing depth and genome coverage of each reference genome were determined with BEDTools coverage v2.27.1 (genomecov -ibam sort.bam -bg)^23^. Reliable co-detection with known respiratory viruses was defined when >50% of the genome was covered.

### Characterization of non-viral microbial communities in hospitalized patients with SARS-CoV-2 infection

Non-viral microbial taxon assignment of the non-human non-rRNA reads was performed using clade-specific marker gene-based MetaPhAln2 with the default parameter options for non-viral microbial composition (╌ignore-viruses)^24^. The presence and relative abundances of non-viral microbial taxa at the phylum, genus and species were estimated (**appendix 2 p 7: Table S6**). Mono-dominance of a given microbial taxa (genus or species) was defined if a taxon had a relative abundance >60% in one sample as suggested by a previous study ^25^. Most of the RNA reads of the two predominant bacterial genera *Burkholderia* and *Parabacteroides*, respectively, identified in the respiratory and gastrointestinal tract of severe cases could hardly be assigned to species level by MetaPhAln2, which might reflect that the two genera contain closely related species that are difficult to differentiate by marker genes. In order to determine which species and how abundant species were in samples mono-dominated by *Burkholderia* or *Parabacteroides*, we downloaded four reference genome sequences of the most frequently isolated BCC species (*B. cenocepacia* J2315, *B. multivorans* ATCC BAA-247, *B. cepacia* ATCC 25416 and *B. dolosa* AU0158) and two gut *Parabacteroides* species (*P. distasonis* ATCC 8503 and *P. merdae* NCTC13052). For each sample, reads were mapped against corresponding references by bowtie2 v2.3.0, and the sequencing depth and genome coverage were estimated by BEDTools coverage v2.27.1 as described above. The summary of coverage and depth of reference genomes for selected samples are presented in **appendix 2 p 8: Table S7**. Likewise, metatranscriptomics-detected bacterial or fungal respiratory co-infections were defined if the respiratory specimens (at least one sample) of severe patients were mono-dominated (relative abundance >60%) by pathogenic microbes known to cause nosocomial infections (**appendix 2 p 9: TableS8**).

### Identification of expressed virulence factors (VFs) in *B. cenocepacia*

To identify the presence and expression patterns of potential virulence factors in *B. cenocepacia* identified in P01, we collected multiple functional categories of virulence genes previously studied and verified by gene mutation analysis in *B. cenocepacia* strains as well as corresponding gene ID in the annotated J2315 genome ^26^, including 1) resistance to stress conditions, 2) antimicrobial resistance, 3) quorum sensing, 4) iron uptake, 5) flagella and cable pilus, 6) lipopolysaccharide and 7) exopolysaccharide. In addition, a pathogenicity island identified on chromosome 2 (BCAM0233-BCAM0281) of *B. cenocepacia* J2315 by using comparative genomics was included ^27^. Non-human non-rRNA microbial reads of all samples from P01 were mapped against the reference genome of *B. cenocepacia* J2315 using bowtie2 v2.3.0 as described above and identified by the gene IDs in the J2315 genome. For each sample, only virulence genes with more than 10 mapped reads were retained. A total of 50 expressed virulence genes were identified in clinical samples collected from P01 and presented in **appendix 2 p 10: TableS9**.

To compare the expression levels between different genes, we performed normalization of target gene expression levels among all detected virulence genes using the following equation:

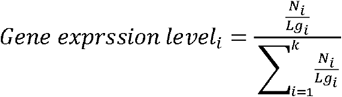

where *i* (1,2,… *k*) refers to a given virulence gene identified in *B. cenocepacia* J2315; *Lg*_*i*_ is the length of gene *i*; *N*_*i*_ is the reads number that mapped to gene *i*.

### Statistical analyses

Non-metric multidimensional scaling (NMDS) ordination of respiratory microbial community was conducted using the Manhattan distances based on a presence/absence matrix of genus profiles of 47 respiratory specimens (7 from mild cases and 40 from severe cases) (R version 3.6.1, vegan package). Kruskal–Wallis test and Wilcoxon rank-sum test were performed to compare differences in SARS-CoV-2 viral loads among five different types of clinical specimens (throat swab, nasal swab, sputum, anal swab and feces) and between overall respiratory specimens from mild and severe cases, respectively (R version 3.6.1, coin package).

## Results

### Demographic information of patients and clinical specimens used in the study

23 patients with COVID-19 hospitalized in the period January 10-March 31, 2020 in four hospitals in the Guangdong Province, China, were enrolled into this study. 15 infected patients (40-80 years) admitted to the ICU and recived mechanical ventilation were defined as having severe COVID-19, and the remaining 8 patients (2-65 years) were mild cases (**appendix 2 p 2: TableS1**). 60 % (9 out of 15) of severely ill patients received invasive mechanical ventilation (**appendix 2 p 2: TableS1**), and 95.7% of the patients (22 out of 23) received antiviral medications. To prevent and control nosocomial infections, all severe cases received broad-spectrum antibiotics, and simultaneously 93.35% (14 out of 15) received antifungal agents **(appendix 2 p 2: TableS1**). By contrast, none of the mild cases were treated with antibacterial or antifugal drugs. Up to March 31, 2020, 53.3% (8 out of 15) of the severely ill patients had been transferred out of ICU or discharged from hospitals, and all mild cases had been discharged, whereas an elderly male patient died one-month after admission to ICU (P01) (**appendix 2 p 2: TableS2**). Sixty-seven serial clinical specimens from the respiratory tract (RT) (n=47, sputum, nasal and throat swab) and gastrointestinal tract (GIT) (n=20, anal swab and feces) of these patients were obtained during the same above period for comprehensive assessment of microbial characteristics after SARS-CoV-2 infection. A detailed timeline of specimen collections and clinical events for the 23 COVID-19 cases are shown in **appendix 2 p 3: TableS2**.

### Work flow of ultra-deep metatranscriptomic sequencing

After quality control, an average of 268.3 Gb metatranscriptomic data were generated per sample (**appendix 2 p 4: TableS3**). We applied an integrated bioinformatics pipeline to detect human, viral and non-viral microbial reads in total RNA-seq data (**appendix 1 p 1: Supplementary figure 1** and **Methods**). The percentage of human RNA reads (including ribosomal RNA and non-rRNA transcripts) varied between different types of specimens, constituting a relatively high fraction of total high-quality reads among RT specimens and a low fraction among GIT specimens (**appendix 1 p 1: Supplementary figure 1** and **appendix 2 p 4: TableS3**). After removing host data, SortMeRNA was applied^21^ to filter microbial rRNA from the metatranscriptomic data. The final remaining non-human non-rRNA data (ranged from 386Mb to 145Gb) were then used to assess viral and non-viral microbial composition by Kraken2X^20^ and MetaPhlan2^24^, respectively(**Methods**). Detailed data statistics for each processing step are provided in **appendix 2 p 4: TableS3**.

### Co-detection of viruses in clinical specimens

First, viral RNA reads belonging to family *Coronaviridae* were defined as SARS-CoV-2-like reads. As expected, *Coronaviridae* was the most abundant virus and was detected in all clinical specimens and varied between 0.01 and 286,418 mapped reads per million (RPM) (**figure 1A**). No statistically significant differences were observed in SARS-CoV-2 viral loads in various types of specimens (**appendix 1 p 2: Supplementary figure 2A**, Kruskal–Wallis test, P=0.561), or respiratory specimens between mild and severe cases (**appendix 1 p 2: Supplementary figure 2B**, Wilcoxon rank-sum test, P=0.455). Although SARS-CoV-2 viral loads in both RT and GIT gradually decreased at later time points of infection (**appendix 1 p 2: Supplementary figure 2C-D**), viral shedding varied in different patients. For instance, RT specimens had consistently lower SARS-CoV-2 viral loads than GIT specimens in P01, while specimens from the two sites showed comparable viral levels in P05 and P10 across all sampled time points (**appendix 1 p 2: Supplementary figure 2E**). Previous studies also reported inconsistent temporal patterns of viral shedding in different types of specimens and patients^28–30^, which might be affected by sample sizes, specimens collected at different stages of infection as well as treatments with a cocktail of antimicrobial agents^31^.

**Figure 1:**
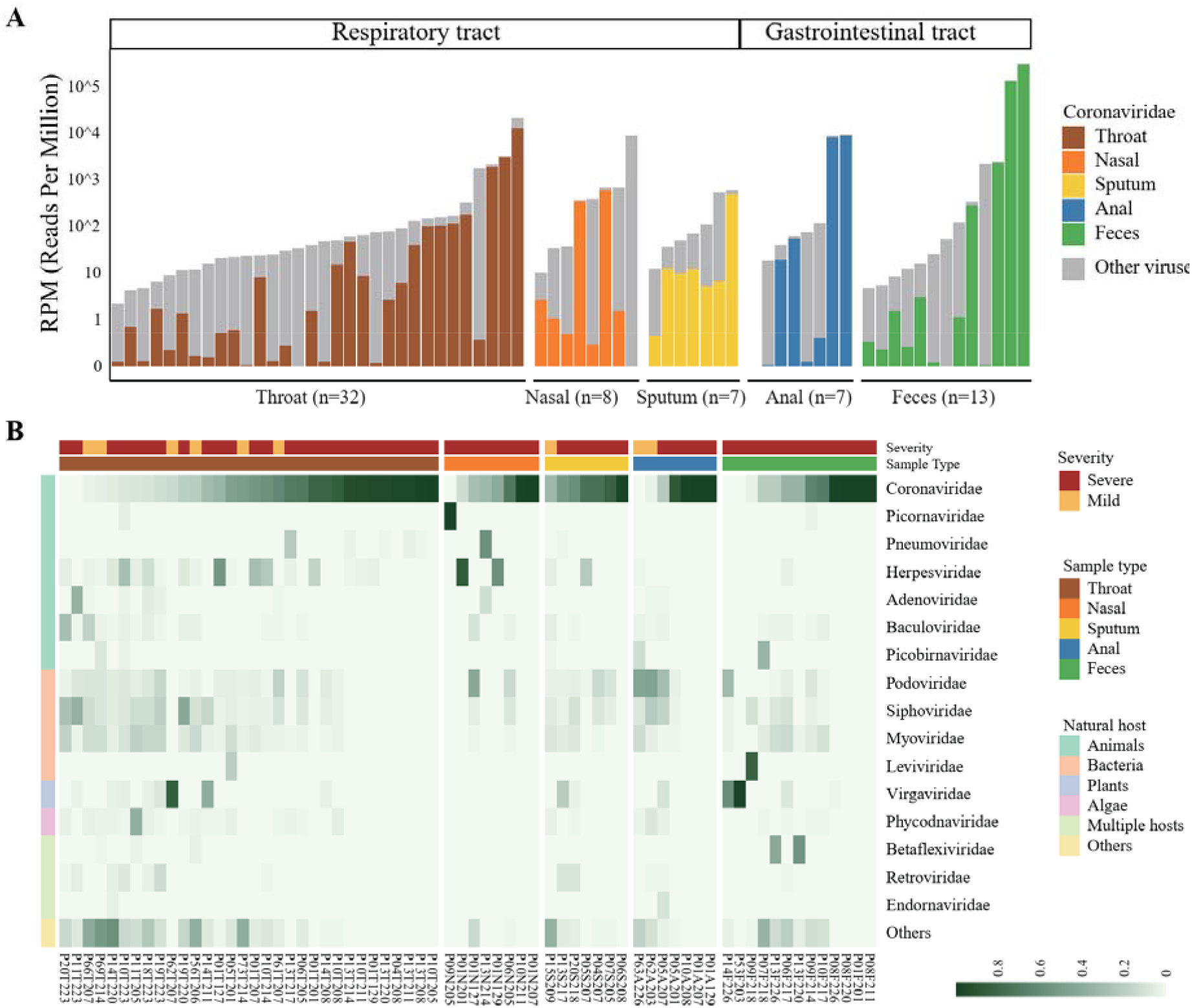
Viral RNA profiles in clinical specimens of hospitalized patients with COVID-19. **(A)** Bar plot showing the number of total viral reads and *Coronaviridae* reads in 67 clinical specimens collected from the respiratory and gastrointestinal tract. Data have been normalized to total sequencing reads in reads-per-million (RPM). The number of *Coronaviridae* reads of different sample types is colored as follows: brown, throat swab; orange, nasal swab; yellow, sputum; blue, anal swab; green, feces. Gray, number of non-*Coronaviridae* viral reads. **(B)** Heatmap showing the viral RNA abundance at the family level. Top16 viral families are shown and ranked according to their natural hosts: green, animals; pink, bacteria; light blue, plant; purple, algae; light green, multiple host species; yellow, others (viral families of low abundances). Specimens from patients with mild and severe COVID-19 symptoms are colored by brick red and orange, respectively.

Except for *Coronaviridae*, RNA-seq analysis also revealed a great diversity of viral composition in clinical samples from infected patients. Natural hosts of the highly abundant viruses differed, including but not limited to animals (e.g., *Picornaviridae, Pneumoviridae* and *Herpesviridae*), bacteria (e.g., *Podoviridae, Siphoviridae* and *Myoviridae*) and plants (*Virgaviridae*) (**figure 1B** and **appendix 2 p 5: Table S4**). The co-infections of known human respiratory viruses were further confirmed (viral genome coverage>50%) in four out of thirteen severely ill patients with metatranscriptomic data of respiratory samples (30.8%), including human alphaherpesvirus 1 in P01 and P05, rhinovirus B in P09, and human orthopneumovirus in P13 (**figure 1B** and **appendix 1 p 4: Supplementary figure 3A-B**). The shedding patterns of human alphaherpesvirus 1 and human orthopneumovirus in throat samples of P01 and P13 were similar to those of *Coronaviridae* in the same patient (**appendix 1 p 4: Supplementary figure 3C-D**). By contrast, co-detection of other respiratory viruses was not observed in mild cases without ICU admission (**appendix 2 p 5: Table S4**). Although some case studies reported the co-infection of SARS-CoV-2 and influenza viruses^32,33^, this was not observed in this Guangdong cohort.

Plant viruses belonging to the family *Virgaviridae*, especially Pepper mild mottle virus (PMMoV) and Tomato mosaic virus (ToMV), were found to be the most dominant viruses in the feces (P53F0203 and P14F226) or in the throat swabs (P62T0207) (**figure 2B** and **appendix 2 p 5: Table S4**). Some pioneer studies have also reported strong evidence supporting the presence of PMMoV and ToMV in human-associated samples^34–36^. All these findings have suggested the presence of plant-human virus transmission. However, the potential pathogenicity of plant viruses in humans or in other vertebrates remains largely unknown^37,38^.

**Figure 2:**
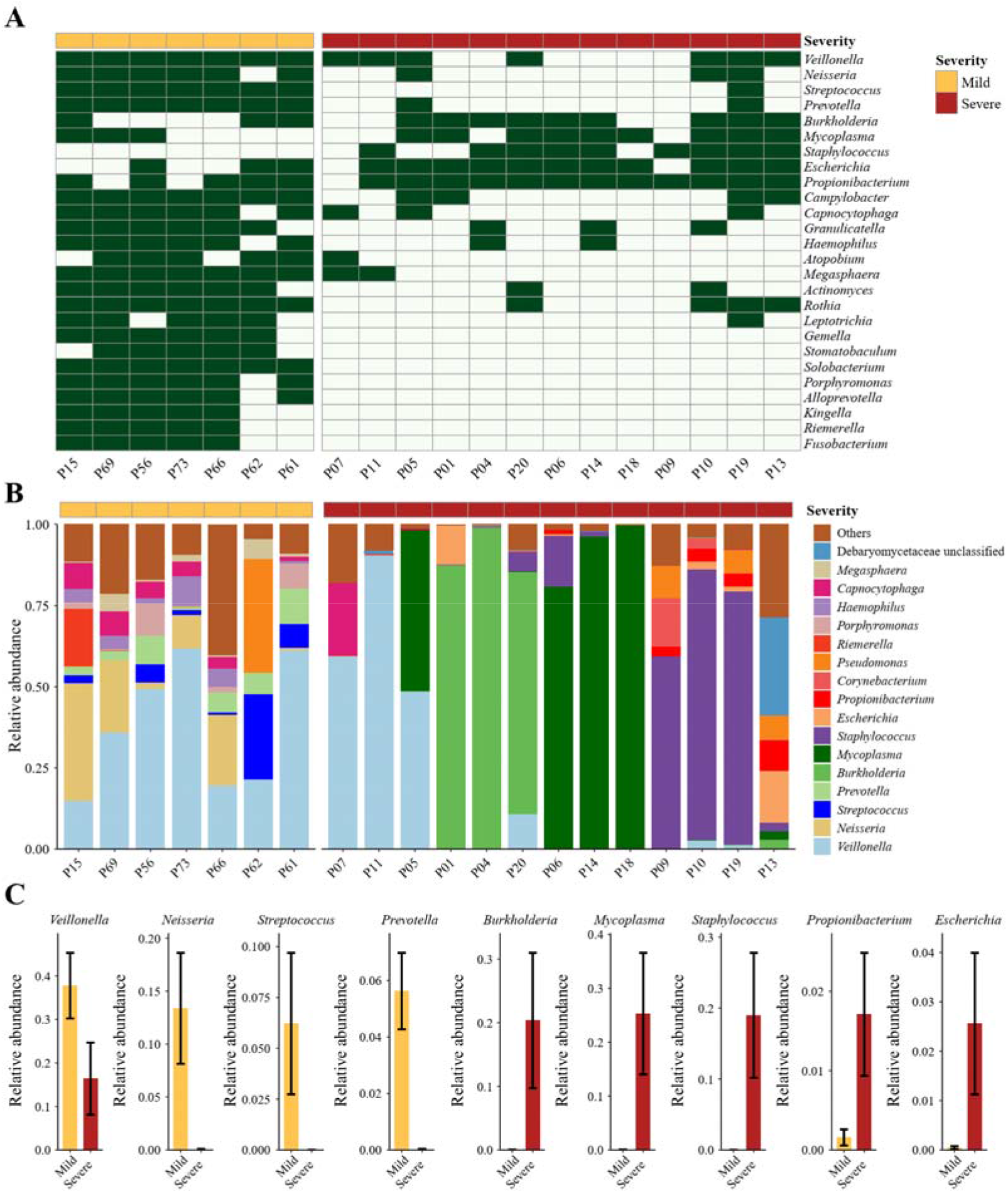
Distinct respiratory microbial signatures in mild and severe cases. **(A)** Presence/absence profile of non-viral microbial genera in mild and severe cases. Orange, mild; brick red, severe. Only common genera detected in over 60% of patients in the mild cases (n>4) or severe cases (n>7) are shown. **(B)** Bar plot showing the relative expression levels of non-viral microbes in mild and severe cases. **(C)** Relative expression levels of selected genera differing between mild and severe cases. For each genus, the bar chart and black error bars denote the mean and standard error values of expression levels in mild (orange) and severe (brick red) cases. For severe cases with more than one sequenced respiratory clinical specimens: the presence of a given genus is considered if any of samples from this patient had positive detection of this genus (relative abundance >0) **(A)**, and the mean relative abundance of each detected genus across all samples is calculated to represent its expression level in this patient **(B-C)**.

### Detection of non-viral microbial co-infections in clinical specimens

We next evaluated the occurrence of bacterial/fungal co-infections that have been shown to be associated with worse clinical outcomes than viral co-infections in hospitalized patients. Notably, results of hospital-laboratory based cultures and assays on specimens demonstrated the presence of nosocomial fungal (n=1) and bacterial infections (n=3) in severely ill COVID-19 patients (**appendix 2 p 2: Table S1**). In detail, one patient (P01) tested positive for (1-3)-β-d-glucan (a common component of the fungal cell wall) in blood samples. Two patients (P04 and P20) had positive sputum cultures for *Burkholderia cepacia* complex (BCC) species, the most common respiratory pathogens causing NP in cystic fibrosis (CF) patients^39,40^. *Staphylococcus epidermidis*, a typical skin bacterium that has been increasingly recognized as a multidrug-resistant nosocomial pathogen^41,42^, was identified by culturing of nasal secretions of one patient (P06).

Next, the non-viral microbial RNA composition of all 67 clinical specimens was analyzed to fully assess possible nosocomial infections. As none of mild cases was admitted to ICU or received antibacterial/antifungal agents, we compared the respiratory microbial communities between mild (n=7) and severe (n=13) cases to determine microbial dysbiosis, co-infection and their associations with clinical outcomes. Remarkable microbial differences in RT specimens between mild and severe cases were observed **(figure 2**). Although the specimens of the mild cases were collected from three different hospitals (Guangzhou, Yangjiang and Qingyuan), their RT samples (six throat swabs and one sputum) consistently exhibited a larger number of detected microbial taxa (**figure 2A** and **appendix 1 p 5: Supplementary figure 4A**) and similar microbial RNA community compositions (**figure 2B** and **appendix 1 p 5: Supplementary figure 4B**). On average, 28 genera and 48 species were detected per RT sample in mild cases while only 6 genera and 4 species were detected per RT sample in severe cases (**appendix 1 p 5: Supplementary figure 4C**). The predominant RT bacteria in mild cases were *Veillonella, Neisseria, Streptococcu*s and *Prevotella* (individual occurrence >80% and mean relative abundance>5%) (**figure 2B**,**C**), which is consistent with common microbial communities found in the human nasal and oral cavity^43^. However, except *Veillonella*, each of the latter three genera enriched in mild cases was only detected in few severe cases (n≤ 3, **figure 2A)** and had a mean abundance of less than 0.05% (**figure 2C**).

Of note, RT microbial features of severe cases were identified to be patient-specific. Among 40 respiratory samples from severe patients, over 60% were mono-dominated (relative abundance>60%) by bacterial genus *Burkholderia* (11 samples from P01, P04 and P20), *Staphylococcus* (6 samples from P10 and P19) or *Mycoplasma* (7 samples from P05, P06, P14, and P18) (**figure 2B**,**C** and **appendix 1 p 5: Supplementary figure 4D**). Each genus was detected in 69.2% of (9 out of 13) severe patients and 92.3% (12 out of 13) of severe patients were positive for at least one of the three genera (**figure 2A**), indicating their prevalence in RT of patients hospitalized with severe COVID-19. By mapping RNA reads to the reference genomes of BCC species (**Methods**), we further confirmed the predominant expression of *B. cenocepacia* in the respiratory tract of P01, and *B. multivorans* in P04 and P20 who had postive sputum cultures of BCC **(appendix 1 p 6: Supplementary figure 5A** and **appendix 2 p 8: Table S7)**. All *Staphylococcus* RNA reads of RT samples from P06 (who also had positive *S. epidermidis* culture), P10 and P19 were also assigned to *S. epidermidis* (**figure 2B** and **appendix 1 p 6: Supplementary figure 5B)**. However, *S. aureus*, a major hospital-acquired pathogen^44^, was not detected in metatranscriptomic data of any of the sequenced RT samples. *Mycoplasma orale* and *M. hominis*, rather than *M. pneumoniae*, were the two identified pathogenic *Mycoplasma* members (**figure 2B** and **appendix 1 p 6: Supplementary figure 5C)**. *Propionibacterium* and *Escherichia* were also frequently detected in RT samples of severe cases (individual occurrence >80%) but were less abundant than the former three genera (mean relative abundance<3%) (**figure 2C**). All the five dominant bacterial genera in severe cases have been frequently associated with nosocomial infections, while they were not detected or present in extremely low abundance in mild cases (relative abundance<0.15%) (**figure 2A**,**C**).

Ascomycetic transcripts (mainly from *Debaryomycetacea*) were identified in all five throat swabs collected from P13 **(figure 2B** and **appendix 1 p 5: Supplementary figure 4D)**. Interestingly, the ascomycetes constituted only 5.7% of the total non-viral microbes at the first time point (8 February) and increased to 18.2% ∼87.4% at later points (11-20 February) (**appendix 1 p 5: Supplementary figure 4D)**. P13, an elderly man in his 70s, had onset of COVID-19 symptoms on January 30 and had been admitted to the ICU since February 5 **(appendix 2 p 2: Table S1)**. These observations suggested a rapid succession from bacteria to fungi had occurred in the respiratory microbiota of P13 three days after ICU admission. These finding indicated that serial monitoring to track other respiratory viruses (human orthopneumovirus, **appendix 1 p 4: Supplementary figure 3**) and secondary fungal infections are required to avoid delayed treatment for such patients. Using the same criterion of microbial monodominance (relative abundance>60%), we determined that a metatranscriptomics-based bacterial co-infection rate among severely ill COVID-19 patients was 69.2% (9 out of 13), and 23.1% (3 out of 13, including P04, P06 and P20) of them had the same results based on laboratory-confirmed bacterial cultures (**appendix 2 p 9: Table S8**). Considering all results from clinical laboratories and metatranscriptomic analyses, we report that 84.6% (11 out of 13) of severely ill patients were co-infected with at least one type of respiratory pathogenic microbes (**appendix 2 p 9: Table S8**).

Additionally, three severe cases (P05, P07 and P11) had high expression levels of *Veillonella* but low levels (or no detection) of the above pathogenic genera in their RT samples **(figure 2A**,**B)**. These individuals had been transferred out of the ICU or discharged from hospitals **(appendix 2 p 2: Table S1)**. Most bacterial pathogens in RT infections, including those we identified in patients with COVID-19, are aerobic or facultative organisms, while *Veillonella spp*. are strictly anaerobic and have been reported to be part of normal oral cavities and rarely isolated in nosocomial infections^45,46^. Of note, all three patients did not receive invasive ventilation during ICU admission (**appendix 2 p 2: Table S1**), and two of them (P07 and P11) were negative for laboratory-confirmed or sequencing-detected co-infections (**appendix 2 p 9: Table S8**).

Similarly, severe cases appeared to have a distinct gut metatranscriptome compared to mild cases. Both of the two mild cases having fecal specimens showed highly abundant *Campylobacter* in the gut metatranscriptome (**appendix 1 p 7: Supplementary figure 6A**), different from previous results of healthy adults^47^. While *Parabacteroides* (including *P. distasonis* and *P. merdae*) mono-dominated the gut microbial transcripts in five samples from four severe cases and displayed a much higher abundance in severe cases than that in mild cases (**appendix 1 p 7: Supplementary figure 6B-D** and **appendix 2 p 8: Table S7**). The extreme bloom of *P. distasonis*, a low abundant but common taxa in the human gut, has been reported after beta-lactam ceftriaxone treatment^25^. Taken together, metatranscriptomic findings have not only complemented and enhanced the laboratory-diagnosed microbial co-infections but also provided comprehensive information of microbial dysbiosis on SARS–CoV-2 infected patients.

### Secondary *B. cenocepacia* infection-associated death of a severely ill COVID-19 patient

In order to identify factors associated with the in-hospital death of patient P01, serial clinical specimens were collected and analyzed. Notably, a distinct timeline for secondary *B. cenocepacia* infection in this patient was clearly observed. On the first day of sampling (January 27 2020), up to 99.9% of non-viral microbial transcripts in his throat swab were assigned to *B. cenocepacia* (P01T127) while most of the transcripts in his nasal swab collected at the same day were from *E*.*coli* (P01N127) (**figure 3A**). *B. cenocepacia* was predominantly present in both throat and nasal swabs for all the following sampling time points (29 January–07 February 2020) (**figure 3A**). In addition, *B. cenocepacia* was detected in all his GIT samples, and the percentage of *B. cenocepacia* to non-viral transcripts gradually increased from 9.5% to 64% (from January 29 to February 07) (**figure 3A)**. The time-dependent dynamics of transcript levels of *B. cenocepacia* suggested that transfer from the upper respiratory tract to the lower gastrointestinal tract had caused a secondary systemic infection in P01. Our findings were also consistent with the patient’s death certificate record indicating septic shock as the cause of death **(appendix 2 p 2: Table S1)**. Indeed, several studies have pointed out that among BCC-infected CF patients, infection with *B. cenocepacia*, rather than other common isolated BCC members (such as *B. multivorans* and *B. cepacia*), constituted the highest risk factor of death^26,40^.

**Figure 3:**
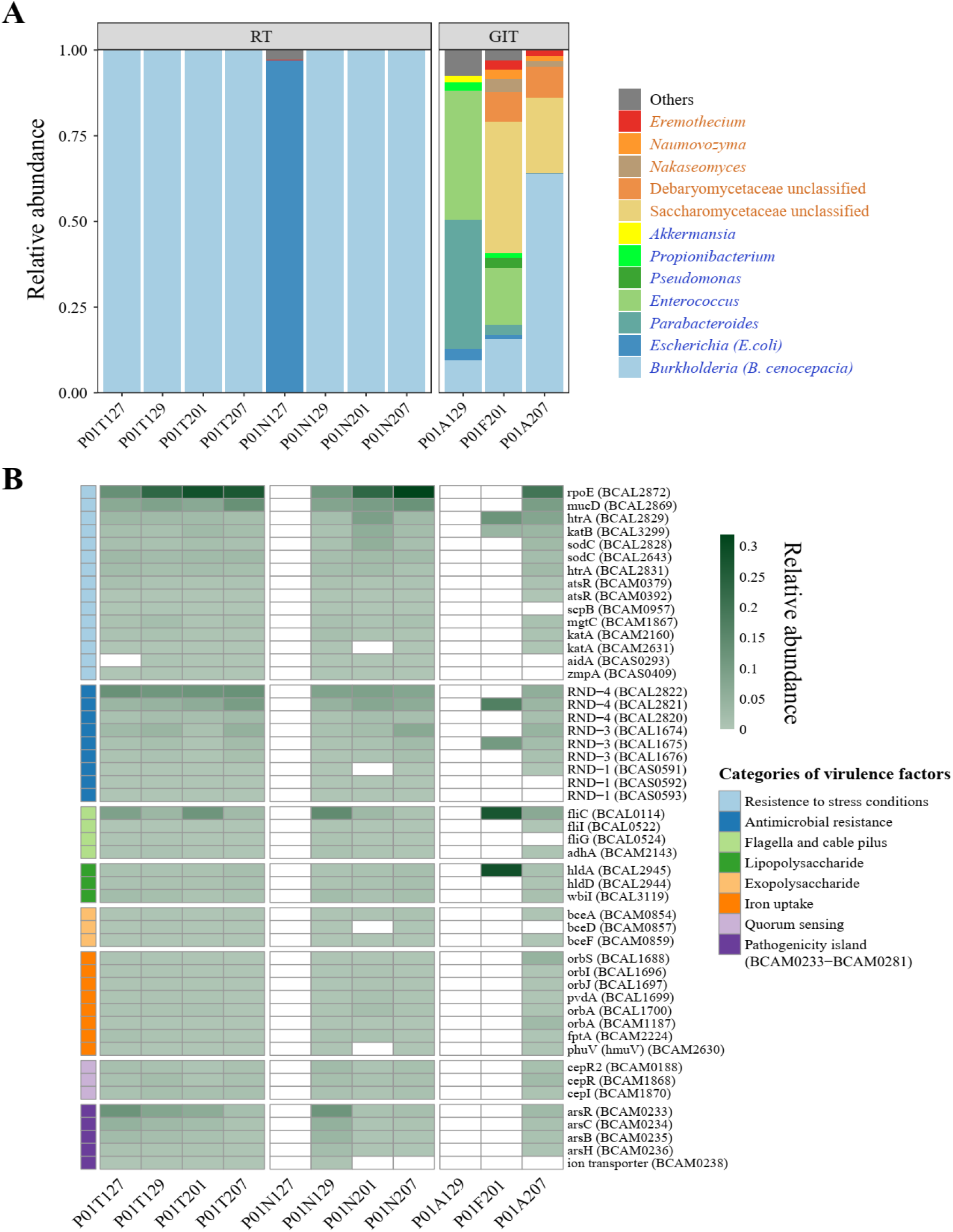
Identification of secondary *B. cenocepacia* infection in P01. **(A)** Bar plot showing the relative expression levels of non-viral microbes in all specimens from P01. A total of 11 specimens from the respiratory tract (RT) and gastrointestinal tract (GIT) are shown. Orange, fungi; blue, bacteria. **(B)** Heatmap showing the relative expression levels of virulence factors of *B. cenocepacia*. A total of 50 identified virulence genes are shown and ranked according to their functional categories: light blue, resistance to stress conditions; blue, antimicrobial resistance; light green, flagella and cable pilus; green, lipopolysaccharide; pink, exopolysaccharide; orange, iron uptake; light purple, quorum sensing; purple, genes located in a pathogenicity island.

Next, virulence factors (VF) expressed by *B. cenocepacia* in P01 were analyzed in order to better understand the pathogenic mechanisms of this possible lethal pathogen in this severe COVID-19 case (**Methods**). The gene *rpoE* (a member of the extracytoplasmic function subfamily of sigma factors) was the most abundantly expressed VF during the entire sampling period in P01 with SARS-CoV-2 infection (**figure 3B)**. RpoE, as a stress response regulator, has been demonstrated to be essential for the growth of *B. cenocepacia* and the delay of phagolysosomal fusion in macrophages during infection^48^. A delay in phagolysosomal fusion has also been reported to be an important host immune escape strategy for several bacterial pathogens^49^. Other VFs in response to oxidative stress conditions in the host environment, such as those encoding superoxide dismutase, peroxidase or catalase (*sodC* and *katB*), were also expressed (**figure 3B, appendix 2 p 10: Table S9)**. A panel of genes belonging to RND (Resistance-Nodulation-Division) family transporters that confer multidrug resistance to *B. cenocepacia* ^50^ were also expressed in large amount in all types of specimens. We also detected expressions of genes encoding flagella and cable pilus (*fliC, flil, fliG* and *adhA*), which can facilitate the bacterial adhesion to host cells and mucin^51^ (**figure 3B**). Besides, expressions of genes involved in quorum sensing, iron uptake (by competing with the host for iron), biosynthesis of lipopolysaccharide (LPS) and exopolysaccharide (EPS) were also detected (**figure 3B**), indicating their active roles in the regulation of bacterial cell aggregation, biofilm formation and toxin production during infection.

## Discussion

In this study, ultra-deep metatranscriptomic sequencing in combination with clinical laboratory diagnosis, including cultures, and colorimetric assays provided a characteristic spectrum of co-detected respiratory pathogenic microbes in a Guangdong hospitalized patient cohort infected with SARS-CoV-2, which is distinct from prior results on the most common bacterial co-infections identified in previous coronavirus outbreak and influenza pandemics^4–8^ or common pathogens associated with nosocomial respiratory infections in China^52,53^.

In particular, respiratory co-infections with BCC were detected in 23.1% of severe cases with evidence from both laboratory cultures and metatranscriptomic results. The serial metatranscriptomic data of all specimens from P01 have revealed the timeline of a secondary infection with *B. cenocepacia* alongside the expression of various virulence genes, which could confer the abilities of the lethal pathogen to evade host defenses (e.g., *rpoE*), adhere target tissues (e.g., flagella-coding genes and *adhA*), produce toxins (e.g. genes encoding biosynthetic enzymes for production of LPS and EPS) and resist the effects of multiple antibiotics (RND family), which eventually may have led to the fetal outcome of the patient. Interestingly, some severely ill COVID-19 patients have been reported to display CF-like pulmonary pathological features including mucus plugs, bronchial wall thickening, and a subsequently impaired mucociliary clearance^54–57^, which could promote the adherence and colonization of mucin-degrading pathogens in the respiratory tract. Furthermore, BCC bacteria, a major threat to hospitalized CF patients, were found to be predominantly localized in the phagocytes and mucus of CF patients^58^. Even though the small sample size in our study limited our ability to assess the general prevalence of BCC infections among severely ill SARS-CoV-2 patients, our findings highlight the need for monitoring and controlling nosocomial BCC/SARS-CoV-2 co-infection by using rapid diagnostic technologies such as PCR-based multilocus sequence typing and enzyme-linked immunosorbent assay. Metatranscriptomic data also revealed that 30.8% of severely ill patients had respiratory tract co-infections associated with *Mycoplasma spp*. (including *M. hominis* and *M. orale*), which lack a cell wall and have inherent resistance to commonly administered beta-lactam antibiotics. Although *Mycoplasma* usually causes mild illness, it has also been associated with serious infections in seniors and immunocompromised individuals^59,60^, who have a high risk for developing severe symptoms from COVID-19. Moreover, we did observe super-high expression levels of *M. orale* in the RT of two severely ill patients (P14 and P18) with prolonged ICU stay (>30 days). Thus, our results suggest that the possibility of *Mycoplasma*-associated co-infection in severely ill COVID-19 patients warrants increased attention.

The SARS-CoV-2 infection is often accompanied by GI symptoms such as vomiting, diarrhea and abdominal pain^61,62^, and the virus has been recently been proven to infect the human gastrointestinal tract ^63–65^. The dominating Proteobacteria transcripts in the gut of antibiotic treatment naïve mild COVID-19 cases in this study as well as the marked gut microbial changes during respiratory viral infections in prediabetes reported in a recent longitudinal multi-omics study^66^ both support interactions between host, respiratory viruses and the commensal microbiota, possibly via the mucosal immune system^67,68^.

Currently, no effective drugs have been licensed for human use against SARS-CoV-2 infection, but a preliminary study has shown an effect of early treatment with Remdesivir^69^. Instead, the widespread use of antimicrobial agents (including broad-spectrum antibiotics) has been documented in many studies (including ours)^9,12^ for the prevention and treatment of secondary infections in COVID-19 patients, especially in those who needed mechanical ventilation. Without initial specimens before any treatments, it is difficult to distinguish to what extent differences in the respiratory microbial patterns associated with SARS-CoV-2 infections reflect the disease or the antimicrobial treatment, or both. Still, our results suggest that the distinct bacterial communities detected in the respiratory and gastrointestinal tract of the severely ill patients might relate to the significant disruption of the normal human microbiota caused by the treatment with antibiotics, allowing colonization by pathogenic antibiotic-resistant bacteria. Worldwide attention should be drawn to potential future threats from a blooming reservoir of antimicrobial-resistant organisms and associated genes after the COVID-19 pandemic.

## Contributors

J.Z., J.L., Y.L and X.X. conceived the project. Y.W., L.Z., Z.Z., A.Z., J. Zhao., W.H., F.X., F.Ye., B.Z. and S.R. collected clinical specimens and clinical information from patients, processed of RNA extraction, and performed in-hospital diagnostic tests. Ji.L. and P.R. performed metatranscriptomic library construction. H.Z., Z.S., H.R., F.Yang., T.L., J.Z. and F.L. processed the sequencing data and conducted bioinformatic analyses. H.Z., Z.S., H.R., and J.L. interpreted the data. H.Z. and J.L. wrote the first version of the manuscript. J.Z., K.K. and H.M.T. contributed substantially to the revisions of the manuscript. All other authors participated in discussions, provided useful comments and revised the manuscript. All authors approved the final version.

## Data Availability

The non-human metatranscriptomic sequencing data from 67 clinical specimens that support the findings of this study have been deposited into CNSA (CNGB Nucleotide Sequence Archive) of CNGBdb with accession number CNP000106 (https://db.cngb.org/cnsa/). Details of software, code and parameters used for data analyses of the current study are provided and publicly available from GitHub (https://github.com/BGI-IORI/RNA-seq-2019nCov).

## Acknowledgments

This work was funded by National Science and Technology Major Project of China (No:2017ZX10303406), the National Major Project for Control and Prevention of Infectious Disease in China (2018ZX10301101-004), the emergency grants for prevention and control of SARS-CoV-2 of Ministry of Science and Technology (2020YFC0841400) and Guangdong province (2020B1111320003, 2020B111108001, 2018B020207013), Guangdong Provincial Key Laboratory of Genome Read and Write (No. 2017B030301011), Guangdong Provincial Academician Workstation of BGI Synthetic Genomics (No. 2017B090904014), and Shenzhen Engineering Laboratory for Innovative Molecular Diagnostics (DRC-SZ[2016]884). Metatranscriptomic sequencing of this work was supported by China National GeneBank.

